# Bladder and bowel management in spinal cord-injured women throughout pregnancy and childbirth: A scoping review protocol

**DOI:** 10.1101/2025.07.07.25331009

**Authors:** Silvia Giagio, Lorenza Maria Landi, Andrea Turolla, Manuela Marani, Maria Giulia Cinotti, Mimosa Balloni, Elisa Mercante, Laura Simoncini

## Abstract

**Background:** Women with spinal cord injury (SCI) may face unique clinical challenges during pregnancy, childbirth, and the postpartum period due to neurogenic bladder and bowel dysfunction. These include urinary retention, incontinence, recurrent urinary tract infections, constipation, along the risk of autonomic dysreflexia. Despite an increasing number of pregnancies in this population, evidence on management strategies remains fragmented. This scoping review aims to map the literature on bladder and bowel conditions in pregnant women with SCI, describe current management practices during pregnancy, childbirth, and the post-partum period, as well as identify any research gaps.

**Methods:** This scoping review will follow Joanna Briggs Institute (JBI) methodology and be reported according to the PRISMA-ScR checklist. Studies will be included if they involve women with SCI diagnosed before conception and report on bladder or bowel conditions or their management during any perinatal phase. All study designs and relevant gray literature will be considered. A comprehensive search strategy will be implemented across MEDLINE, EMBASE, Scopus, and Cochrane databases, without date restrictions, including articles in English and Italian. Study selection and data extraction will be conducted independently by two reviewers. Data will be presented numerically and thematically, with subgroup analysis by perinatal phases, medical conditions, type and level of SCI, and clinical management strategy.

**Conclusions:** This review will provide a comprehensive synthesis of existing evidence on bladder and bowel management in pregnant women with SCI. Findings will inform clinical practice and guide future research, contributing to the development of evidence-based care pathways for this population.

## INTRODUCTION

Individuals with spinal cord injury (SCI) are at risk of a wide range of medical complications including autonomic dysfunction, neurogenic bowel and bladder, pressure ulcers, venous thromboembolism, osteoporosis, and muscle spasticity [1,2].

Among women, SCI does not impair fertility [3,4] and although pregnancies remain relatively uncommon, they are on the rise and generally yield favorable outcomes [3–5]. However, the physiological changes of pregnancy can exacerbate pre-existing conditions such as urinary retention, incontinence, and the risk of recurrent urinary tract infections (UTIs) [2]. Additionally, neurogenic bowel, often manifesting as constipation, can precipitate autonomic dysreflexia (AD), a potentially life-threatening condition [6]. Given these complexities, healthcare providers require evidence-based guidance for managing these conditions, yet research in this area remains fragmented.

This scoping review therefore will aim to systematically map the existing literature on bladder and bowel management in pregnant women with SCI, identify best practices, and highlight knowledge gaps in research.

### RESEARCH QUESTION(S)

1. What bladder and bowel medical conditions may affect women with spinal cord injury during pregnancy, childbirth, and the postpartum period?
2. What are the current clinical practices and recommendations for bladder and bowel management in SCI women during pregnancy, childbirth, and the post-partum period?
3. How are management strategies adapted according to the clinical stage (pre-partum, labor, post-partum), level of spinal cord lesion, and completeness of the injury?
4. What are the existing gaps in the literature, and what research priorities emerge to guide the development of evidence-informed clinical practice in this area?

### Inclusion criteria

Studies will be included if they meet the following Population, Concept, and Context (PCC) criteria.

#### Population

Women of any age, at any stage of pregnancy or postpartum, irrespective of the level of the spinal cord lesion or the grade on the ASIA Impairment Scale. Only studies involving women with a diagnosis of SCI established prior to conception will be included. Cases where SCI occurs as a complication during pregnancy or childbirth will be excluded.

#### Concept

Any bladder and bowel condition according to international terminology, as described by authors of studies, including but not limited to urinary retention, incontinence, urinary tract infections UTIs, renal complications and constipation. Any clinical management strategy for bladder and bowel conditions (e.g. conservative, pharmacological, surgical) will be considered. Additionally, studies investigating healthcare professionals’ perspectives on clinical management strategies for bladder and bowel conditions will also be considered for inclusion.

#### Context

Any healthcare setting and geographical area.

### Types of sources

This scoping review will include any primary and secondary research as well as text, book chapters, and expert opinion papers relevant to the clinical management of pregnant women with SCI.

## METHODS

### Study design

This protocol follows best practice guidance for scoping reviews and is developed in accordance with the Joanna Briggs Institute (JBI) methodology [7]. The final scoping review will be reported according to the Preferred Reporting Items for Systematic Reviews and Meta-Analyses extension for Scoping Reviews (PRISMA-ScR) Checklist [8].

### Research team

The review will be conducted by a multidisciplinary team with expertise in research methods, pelvic floor dysfunction, urogynecology, physical medicine and rehabilitation, and neurorehabilitation.

### Search strategy

An initial limited search of MEDLINE was undertaken to identify articles on the topic. The text words contained in the titles and abstracts of relevant articles, and the index terms used to describe the articles were used to develop search strategies for MEDLINE (**see Appendix 1**). The databases to be searched include Medline, EMBASE, Scopus, Cochrane. Additionally, gray literature sources will be searched. The reference list of all included sources of evidence will be screened for additional studies. Studies published in English and Italian language will be included. No time limit will be imposed on the papers for inclusion in this review.

### Source of evidence selection

The study selection process will consist of two levels of screening using Rayyan: (1) a title and abstract and (2) full-text review. For both of levels, two authors (SG, MM) will independently screen the articles with conflicts resolved by a third author. Reasons for the exclusion of any full-text source of evidence will be recorded and presented in the latest published version of the Preferred Reporting Items for Systematic Reviews and Meta-analyses (PRISMA) flow diagram.

### Data extraction

Data will be extracted into a Microsoft Excel template by two independent reviewers (SG, MM). Main information including study characteristics, medical conditions, stages of pregnancy, management strategies, healthcare settings will be extracted.

A draft extraction form is provided (see **Appendix 2**). Charting results is commonly an iterative process during scoping reviews; other data can be added to this form according to the subgroups that could emerge from the analysis of the studies included. Modifications will be detailed in the full scoping review. Any disagreements that arise between the reviewers will be resolved through discussion. If appropriate, authors of papers will be contacted to request missing or additional data, where required.

### Critical appraisal of individual sources of evidence

Formal quality assessment will not be conducted, as per scoping review methodology [7].

### Data analysis and presentation

The results will be presented in two ways:

1. Numerically. Studies identified and included will be reported, and the description of the search decision process will be mapped. In addition, extracted data will be summarized in tabular and diagrammatic form.
2. Thematically. A thematic summary will be performed pertaining to themes and key concepts relevant to the research questions and according to subgroups (e.g. medical condition, stage of pregnancy, categorization of management strategies) and to others that could emerge. Finally, a synthesis of the reports will highlight the research gaps.

This approach will enable the creation of a comprehensive framework for management.

### Dissemination

Findings will be published in peer-reviewed journals, presented at international conferences, and shared with medical associations specializing in urogynecology, physical medicine and rehabilitation, and neuro-urology.

## Data Availability

All data produced in the present study are available upon reasonable request to the authors.

## Acknowledgements

Not applicable.

## APPENDIX

Appendix 1: Preliminary search strategy (MEDLINE)

Appendix 2: Draft data extraction instrument

### Appendix 1

Search conducted: April 27, 2025

| MEDLINE | RESULTS |
| --- | --- |
| ("Spinal Cord Injuries"[MeSH Terms] OR "Spinal Cord"[Title/Abstract] OR "SCI"[Title/Abstract] OR "spinal injury"[Title/Abstract] OR "spinal trauma"[Title/Abstract] OR "paraplegia"[MeSH Terms] OR "paraplegia"[Text Word] OR "tetraplegia"[Text Word] OR "quadriplegia"[Text Word] OR ("neurogenic"[Title/Abstract] AND "bladder"[Title/Abstract]) OR ("neurogenic"[Title/Abstract] AND "bowel"[Title/Abstract]) OR "neurogenic bowel"[MeSH Terms] OR "urinary bladder, neurogenic"[MeSH Terms]) AND ("Pregnancy"[MeSH Terms] OR "Pregnant Women"[MeSH Terms] OR "pregn*"[Text Word] OR "antenatal"[Title/Abstract] OR "prenatal"[Title/Abstract]) | 4,342 |

### Appendix 2

#### For primary studies

|  |  |
| --- | --- |
| **Author(s)** |  |
| **Aim(s)** |  |
| **Study characteristics** | Year of publication, study design |
| **Population characteristics** | Number of participants, age |
|  | Type of SCI (e.g. traumatic, non-traumatic), level of lesion, ASIA Impairment Scale, time since injury |
| **Clinical condition of interest** | Urinary and bowel conditions as authors described (e.g. UTI, constipation)<br>Stage of pregnancy (pre-partum, labor, post-partum) |
| **Other medical condition** | Any other medical diagnosis |
| **Clinical management** | Management strategies used or discussed including but not limited to: conservative (e.g. catheterization, rehabilitation and physical therapy, other conservative approaches), pharmacological, surgical. Procedures description, including clinical recommendations. Follow up. |
| **Context** | Country, healthcare setting, healthcare providers involved (e.g. medical physicians, healthcare professionals) |
| **Outcomes and findings** | Clinical outcomes (symptom improvement, incontinence episodes, urinary tract infections), complications reported, quality of life outcomes, patient-reported experiences and preferences |
| **Additional notes** | Key conclusions, identified gaps in research, other relevant findings |

### For secondary studies

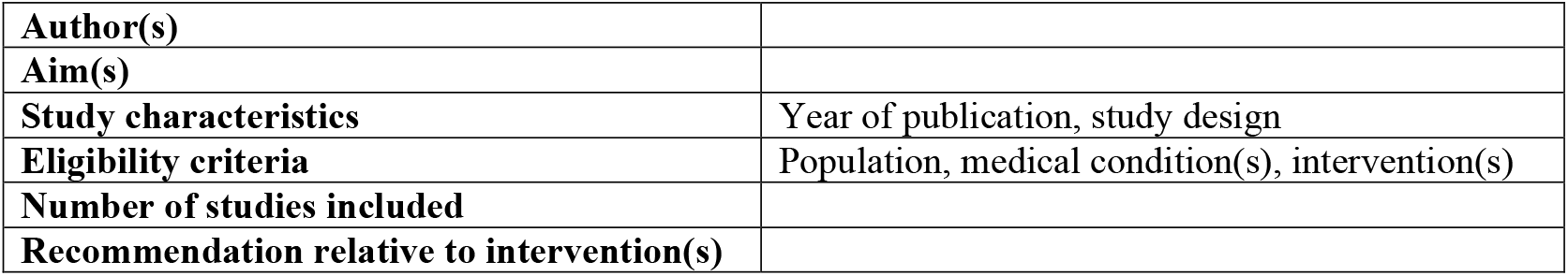

|  |  |
| --- | --- |
| **Author(s)** |  |
| **Aim(s)** |  |
| **Study characteristics** | Year of publication, study design |
| **Eligibility criteria** | Population, medical condition(s), intervention(s) |
| **Number of studies included** |  |
| **Recommendation relative to intervention(s)** |  |

#### For recommendations and guidelines

|  |
| --- |
| **Author(s)** |
| **Year of publication** |
| **Organism or state implicated** |
| **Country of origin** |
| **Recommendation relative to intervention(s)** |

